# Mapping the context of sedentary behaviour (MAPS-B) using wearable sensors, indoor positioning systems, and diaries in older adults who are pre-frail and frail: A feasibility longitudinal study

**DOI:** 10.1101/2023.08.04.23293656

**Authors:** Isabel B. Rodrigues, Rachel Swance, Suleman Tariq, Alexa Kouroukis, Jonathan Adachi, Steven Bray, Alexander Rabinovich, George Ioannidis, Rong Zheng, Dylan Kobsar, Qiyin Fang, Alexandra Papaioannou

**Author notes:** Corresponding author (IBR). Department of Medicine, University of McMaster, Hamilton, Ontario, Canada.

## Abstract

Older adults who are frail are likely to be sedentary. Prior interventions to reduce sedentary time in older adults have not been successful as there is little research about the context of sedentary behaviour (posture, location, purpose, social environment). Moreover, there is limited evidence on feasible measures to assess context of sedentary behaviour in older adults. The aim of our study was to determine the feasibility of measuring context of sedentary behaviour in older adults with pre-frailty or frailty using a combination of objective and self-report measures. We defined “*feasibility process”* using recruitment (20 participants within two-months), retention (85%), and refusal (20%) rates and “*feasibility resource*” if the measures capture context and can be linked (e.g., sitting-kitchen-eating-alone) and are all participants willing to use the measures. Context was assessed using a wearable sensor to assess posture, a smart home monitoring system for location, and an electronic or hard-copy diary for purpose and social context over three days in winter and spring. We approached 80 potential individuals, and 58 expressed interest; of the 58 individuals, 37 did not enroll due to lack of interest or medical mistrust (64% refusal). We recruited 21 older adults (72±7.3 years, 13 females, 13 frail) within two months and experienced two dropouts due to medical mistrust or worsening health (90% retention). The wearable sensor, indoor positioning system, and electronic diary accurately captured one domain of context, but the hard copy was often not completed with enough detail, so it was challenging to link it to the other devices. Although not all participants were willing to use the wearable sensor, indoor positioning system, or electronic diary, we were able to triage the measures of those who did. Future studies will need to determine the most feasible and valid method to assess the context of sedentary behaviour, especially in diverse older adults.

## Introduction

Older adults who are frail are more likely to be sedentary (1,2). Frailty is a multidimensional syndrome characterized by a decline in function across multiple physiological systems including the cardiovascular, musculoskeletal, neurological, and immunological systems (3,4). Sedentary behaviour is defined as any activity during awake hours in a seating, reclining, or laying posture that uses low energy expenditure (i.e., ≤ 1.5 metabolic equivalent of task [MET]) (5). Sedentary behaviour is not merely the absence of moderate or vigorous physical activity but also a reduction in sit-to-stand transitions, stand time, and light physical activity (6). Most older adults who are frail spend 60% of their awake time in a seated or laid position (6). Prolonged periods of sedentary time can lead to muscle and bone unloading and are associated with declines in mobility and quality of life, and increased risk of falls, fractures, and death (1,6–9). In addition, prolonged screen-based sedentary activities are associated with both depressive and metabolic syndromes (2). The deleterious health effects of sedentary behaviour are different to those of physical inactivity and are partially independent of an individual’s physical activity levels (6). Even older adults who meet the recommended aerobic exercise guidelines of moderate to vigorous physical activity might experience adverse effects of sedentary behaviour (6). Thus, interventions to reduce periods of prolonged sedentary behaviour are necessary, especially among older adults who are living with frailty.

Prolonged sedentary behaviours are a recognised risk factor for many medical disorders, which makes it an urgent objective for preventative health interventions. To evaluate the effectiveness of such interventions, measures that are responsive to change are required (10). Although accelerometer-derived assessments indicate that older adults have the highest levels of sedentary time (11), these objective measures do not provide contextual information to identify interventions or public health messages to reduce sedentary time (10,12). Inclinometers are the most sensitive and valid measure of total sedentary time, but the limitation of such devices is its inability to accurately assess specific modalities of sedentary behaviour (13) Moreover, device-based measures have a high cost-to-utility ratio, which often limits their use in research (14). A recent meta-analysis reported that current tools for assessing context of sedentary behaviour or total sedentary time either over-report or under-report the amount of time adults and older adults spend sitting (12). For example, single item self-report questionnaires typically underestimate sedentary time when compared to device-based measures (accelerometers and inclinometers) (12). On the other hand, multi-item questionnaires, ecological momentary assessments, and diaries with a short recall period are more accurate at measuring sedentary time; however, there is also a high degree of variability between and within those tools (12). Currently, there is no gold standard to assess the context of sedentary behaviour, especially in older adults.

Almost all studies in older adults have assessed total sedentary time, which does not provide enough information to understand the context of sedentary behaviours (2,8). The main reason to understand context is because not all sedentary behaviours need to be modified as some cognitively engaging sedentary behaviours (e.g., reading, socializing) appear to benefit health, while time spent in more passive activities may be detrimental. A sedentary behaviour research priorities international consensus statement suggests researchers should explore objective and self-report methods to assess context of sedentary behaviour among older adults (8). We used the Sedentary Behaviours International Taxonomy to guide our definition of context of sedentary behaviour (15). Context was defined as the purpose of the sedentary behaviours, the location where the behaviours occur, posture of the behaviours (e.g., lying, sitting), social context (e.g., alone or with others), and time of day the behaviours occur. To map the context of sedentary behaviour we used objective (i.e., accelerometer and home monitoring system with an indoor positioning system), and self-report (i.e., diary) measures; we chose three measures as one measure alone does not provide enough information about context. Our study is unique because it uses a combination of measures to assess context of sedentary behaviour; however, the feasibility of these combined measures in older adults is unknown. The primary purpose of our study was to determine the feasibility of using three measures to assess the context of sedentary behaviour in older adults who are pre-frail and frail. Our secondary objectives to quantify the context of sedentary behaviours and to understand perspectives of sedentary behaviour are reported elsewhere.

## Materials and Methods

### Study Design

We conducted a mixed-methods longitudinal study with older adults who are pre-frail and frail. We followed the STROBE 2007 guidelines for reporting of observational studies (S1 Table) (16). Ethics approval was obtained from the Hamilton Integrated Research Ethics Board. We registered our study with clinicaltrials.gov (NCT05661058) on December 22^nd^, 2022. We assessed feasibility over three days (one weekend and two weekdays) in the winter and spring as sedentary behaviour may differ by the season.

### Setting

We recruited participants from physicians’ offices, the local newspaper, and a local radio station. We also posted advertisements on social media using Facebook and Twitter. To ensure diversity in our recruitment process we partnered with CityHousing Hamilton Corporation, an organization that provides subsidized housing to low-income older adults, many of whom are of visible minorities, immigrants, and have visible disabilities (i.e., use a walker or cane). The results of our recruitment and retention strategy of diverse (members of racial and ethnic minorities, diverse genders, low socioeconomic status) are described elsewhere (17). We recruited participants between January to February 2023. We obtained written informed consent from each participant prior to enrolling them in the study. Participants attended two study visits (once in the winter and another in the spring) in a private room at St. Peter’s Hospital, which is part of the Hamilton Health Sciences. We provided free transportation for participants with limited mobility or free parking at the hospital. Participants were grouped into four cohorts of five participants. During the first week, we met with five participants at St. Peter’s where they completed a series of questionnaires and physical performance measures. We provided each participant with a wearable sensor, explained how to set up and calibrate the indoor positioning system, and complete the electronic or hard copy diary. During the second week, we collected the devices and diaries and transferred the data to a McMaster University cloud, and cleaned and charged the devices. We repeated the process with each cohort and the entire process was repeated in the spring. Participants with limited mobility or transportation were provided with pre-paid boxes to return study items. At the end of the study, participants received remuneration as a gift card to an easily accessible location on the bus route with versatile buying options (e.g., groceries, clothing, furniture, cleaning supplies).

### Participants

We included participants if they: 1) spoke English or attended with a translator or caregiver; 2) were ≥60 years and older; and 3) had a Morley Frail Scale score ≥3 (i.e., a score of 0 is robust, a score 1 or 2, pre-frail, and a score of 3 to 5, frail) (18). We excluded individuals who: 1) used a wheelchair for at least 55% of the awake day due to medical conditions; 2) were not independently mobile (i.e., require assistance from another individual to ambulate); and 3) had travel plans or other commitments that required missing >30% of the rollout period. We sought to enroll both males and females as we anticipated that gender may influence sedentary behaviour through socially constructed norms and roles and can be affected by differential access to resources, opportunities, and power.

### Measures and Data Sources

To map the context of sedentary behaviour we used objective (wearable sensor and indoor positioning system), and self-report (daily diary) measures. Participants were equipped with the wearable sensor and indoor positioning system, and completed a diary of daily activities over three days (one weekend and two weekdays) in the winter (February 1, 2023 to March 21^st^, 2023) and spring (April 10^th^, 2023 to May 27^th^, 2023). The three measures were linked using date and time (e.g., sitting-living room-watching TV-alone weekend, Winter 3:30 pm to 5:15 pm).

### Wearable sensor

We used the activPAL4^TM^ to collect data on posture. The activPAL4^TM^ is a valid tool to use among older adults that generates totals for the time spent lying, sitting, standing, and stepping every second of the day (19). The wearable sensor was secured to participant’s right upper thigh, midway between the iliac crest and the upper line of the patella, using a waterproof 3M Tegaderm Transparent bandage. Participants were asked to continue their normal daily activities as the wearable sensor would not interfere with their daily lives. Data was collected on the device’s hard drive and exported manually to a secure McMaster University cloud.

### Indoor Positioning System

We used a custom-designed and developed indoor positioning system to obtain room level positioning information. The system was designed and validated to be used by older adults in their own homes without the need for a floor plan and only minimal initial setup and calibration; the system can also be used in homes with multiple stories with multiple residents (20). The indoor positioning system consists of a smartwatch, a few beacons, and a data hub. The participants wore a commercially available, off-the-shelf smartwatch with customized software. The smartwatches were waterproof and could be used in the shower and pool. The location of the smartwatches is tracked by ambient (nonwearable) beacons plugged in regular wall outlets of different functional rooms of the participant’s homes; we defined functional rooms as areas that participants used at least 25% of the day (e.g., kitchen, bedroom, living room). The system detected location and tracked the room-to-room movements of the participants at seconds intervals (20). The data was collected wirelessly by a data hub and stored on a secured McMaster cloud data server.

### Diary

Each participant was asked to complete a diary of 24-hour daily activities using an electronic diary (Activities Collected over Time over 24-hours (ACT24)) or a hard copy version that asked participants to describe their activity, who they did the activity with, and the date and time. ACT24 was developed by the National Cancer Institute for research purposes (21,22). ACT24 is an internet-based previous-day recall designed to estimate total time (hours/day) spent sleeping in bed, in sedentary behaviours during awake hours, and in physical activity (21,22). ACT24 also provides estimates of energy expenditure associated with each behaviour (MET-hours/day) (21,22). We provided all participants with several sheets of the hard copy diary and participants who used the electronic diary inputted their activities the next day into ACT24. We sent daily email reminders to participants to complete their electronic diary.

### Health outcomes

We collected baseline data on falls in the last 6-weeks, cognition score, frailty status, activities of daily living, health-related quality of life, depression, and anxiety in the winter and spring. Fall history was assessed by asking the following question: “*we would like to know about any falls you have had in the last 6-weeks. Have you had any fall including a slip or trip in which you lost your balance and part or all of your body landed on the floor or ground or lower level*?” (23). Cognitive status was assessed using the Montreal Cognitive Assessment (MoCA); we administered version 8.2 English in winter and MoCA Basic in spring (24). MoCA scores were adjusted for age and education level. Frailty scores were measured using the Fit-Frailty Assessment & Management Application (pre-frail scores 0.18 to 0.24 and frail >0.24) (25), activities of daily living with the Nottingham Extended Activities of Daily Living Scale (26), and health-related quality of life using the EuroQol 5-Dimension 5-Level (EQ-5D-5L) questionnaire (27). We assessed depression scores using the Geriatric Depression Scale (28) and anxiety using the Geriatric Anxiety Scale (GAS-10) (29). Demographic characteristics were collected using PROGRESS (Place of residence, Race/ethnicity, Occupation, Gender and sex, Religion, Education, Socioeconomic status, and Social capital) (30).

### Sample Size

As the primary outcome is feasibility, we selected a sample size of 20, which was considered large enough to understand the practicability of using this novel approach to mapping sedentary behaviour. Sample sizes between 12 to 24 are considered reasonable for feasibility and pilot studies (31,32).

### Outcomes

Our primary outcome was feasibility, which was defined using “feasibility process” and “feasibility resource” (33). Feasibility process included recruitment, retention, and refusal rates, while feasibility resource was determined using the following questions: 1) can each measure capture its intended domain of context (e.g., does the diary capture purpose and social context); 2) can data be triaged by date and time; and 3) are all participants willing to use or complete the measures. Our criteria for success for feasibility process were to recruit 20 participants within two-months with 85% retention and 20% refusal rates. Our recruitment criterion is based on previous frailty research in which 1-in-5 individuals who are approached in clinic are successfully recruited (23,34). We anticipated that the physicians could approach 10 potential participants per week for 8 weeks (80 total participants). Our criteria for retention and refusal rates were based on a frailty systematic review where retention rates range from 70% to 90% and refusal rates from 10% to 20% (35). Our criteria for success for feasibility resources were determined if each measure could capture a domain of context, where if “yes” than feasibility is achieved, while if “no” or “sometimes”, feasibility is not achieved (33). The same dichotomous methods were applied if the measures could be triaged using date and time (yes or no/sometimes), and if participants were willing to use activPAL4^TM^, the indoor positioning system, and complete the ACT24 (yes or no/sometimes for each measure). We also conducted exit interviews with each participant to ask about experiences using each measure.

### Statistical Analysis

Demographic data, feasibility process, and feasibility resources were reported using means and standard deviations or as a count and percentage. Descriptive analyses were performed using Microsoft Excel (version 16.71). Each exit interview was audio-recorded, transcribed verbatim, and analysed using content analysis (36). Missing values were reported as missing. Individuals who were loss to follow-up were included in the analysis if their data were available. Adverse events were reported using narrative description.

## Results

### Feasibility Process

We approached 80 individuals, and 58 expressed interest in the study (Fig 1). Of the 58 individuals, 37 declined (64% refusal rate) to enroll citing lack of interest because they initially thought the study was an exercise trial or they changed their mind (57%), medical mistrust (27%), or worsening medical health (16%). Twenty-eight of the 37 individuals who declined to participate identified as female, 1 as transgender male, and all 37 individuals had a Morley frail score ≥ 3. We enrolled 21 participants within two months. About 71% of participants were recruited from a physician’s office, 19% from advertisements posted in CityHousing Hamilton, and the other 10% from community advertisements posted on social media or the radio. A day after the initial study visit, one participant withdrew citing medical mistrust in the study and another participant withdrew after completing the winter period citing worsening medical health (90% retention rate). Both individuals who were lost to follow up were over the age of 75 years and categorized as frail. Five participants required transportation and three utilized the pre-paid box option.

**Fig 1.**
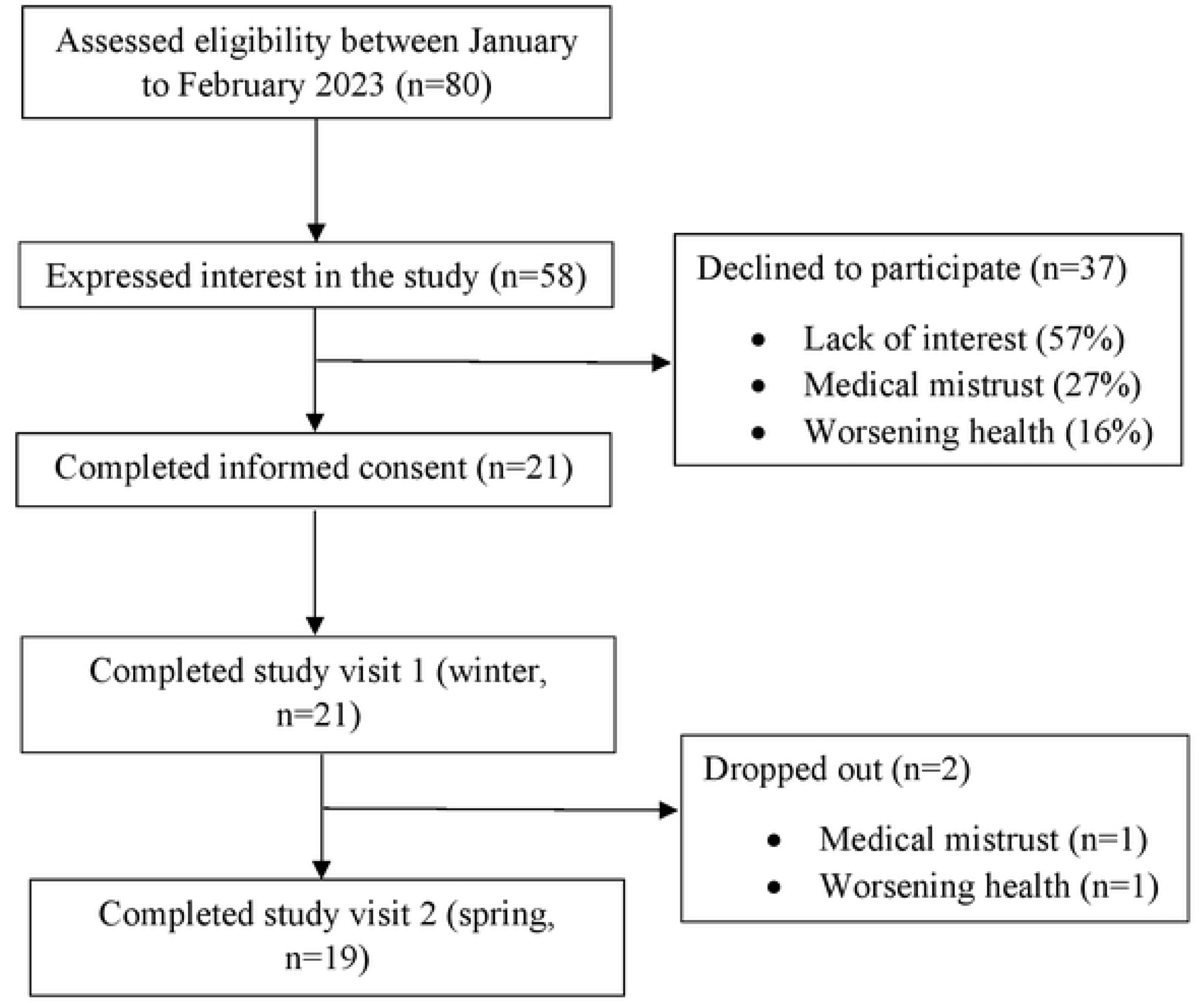
Flow diagram of recruitment and retention process

### Participant Characteristics

Baseline characteristics are presented in Table 1. Participants who were frail had poorer scores on the Geriatric Depression Scale, Geriatric Anxiety Scale, gait speed, and 5x sit-to-stand compared to individuals who were pre-frail. There were no differences between the frail and pre-frail group on the MoCA, grip strength, EQ-5D-5L, and the Nottingham Activities of Daily Living. We also found no differences between physical performance measures and health outcomes between the winter and spring. One participant did not complete the MoCA in winter because they forgot their reading glasses, and another did not complete the Geriatric Depression Scale in spring for personal reasons.

**Table 1:**
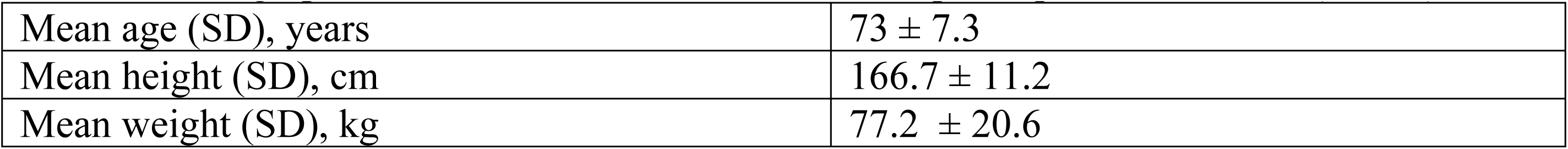
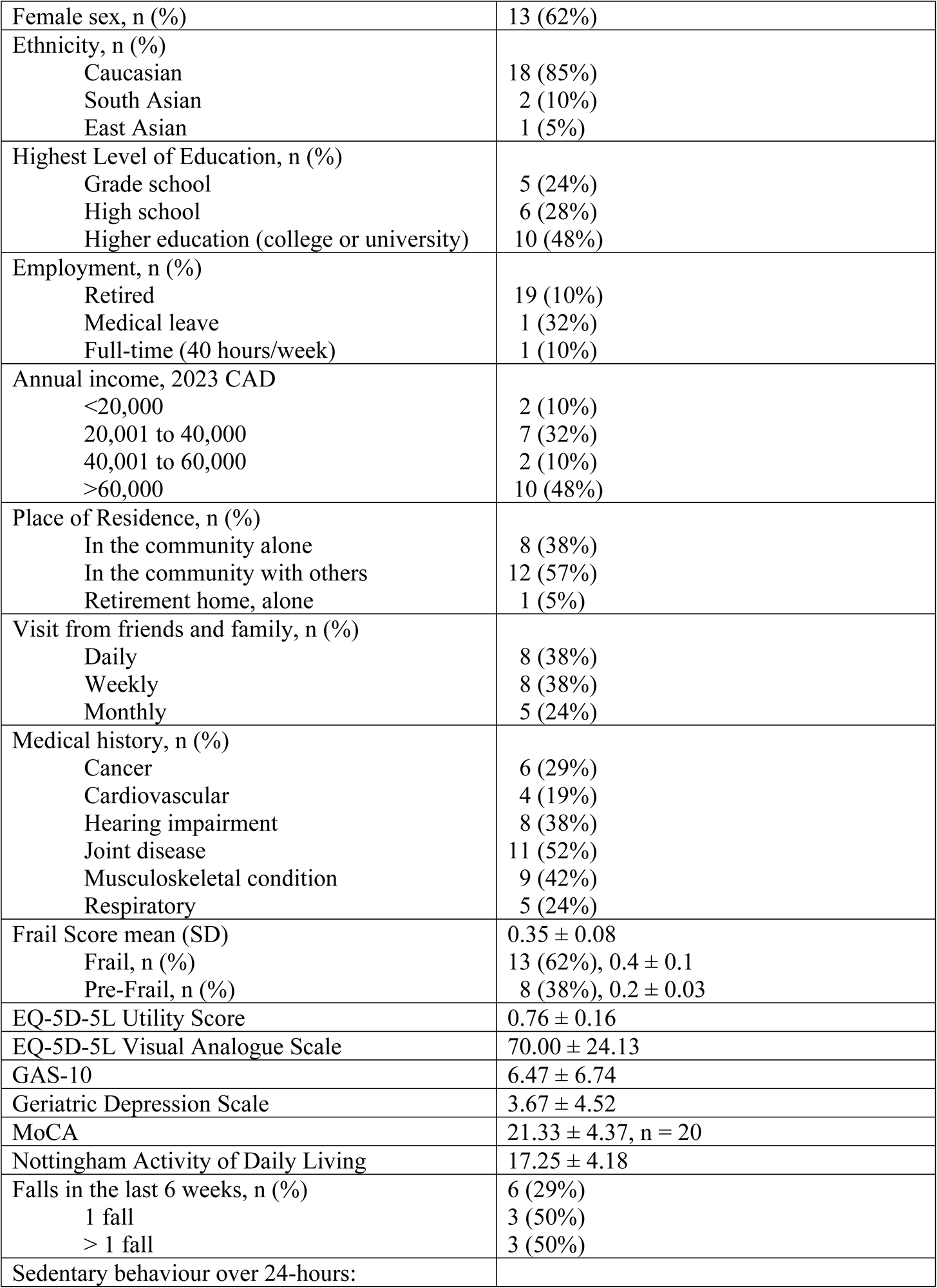
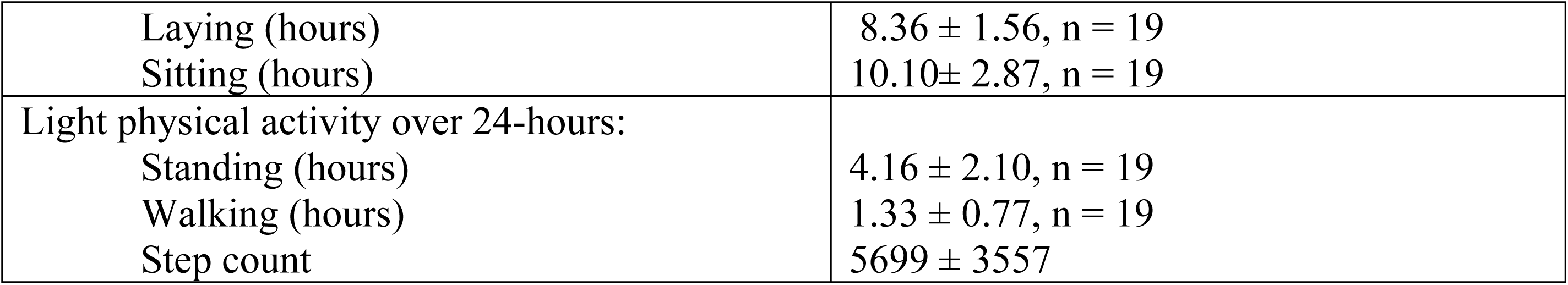
Demographic and other health characteristics of participants at baseline (n = 21)

### Feasibility Resource

We found that each measure captured its domain of context except for the hard copy diary. The hard copy diary was not completed with sufficient information about the activity or time of day. We were able to link data from activPAL4^TM^, the indoor positioning system, and ACT24, but not with the hard copy diary. All participants were willing to complete the hard copy diary while almost all participants were willing to use the wearable sensor. Only some participants opted to use the indoor positioning system and complete ACT24.

#### Wearable sensor

Twenty of the 21 participants felt comfortable using the activPAL4^TM^ device to assess posture. One participant initially agreed to wear the device, and then removed it immediately after the study visit citing medical mistrust. All 20 participants found the device “comfortable to wear” and that they “did not really notice it”. Participants wore the devices continuously for three or four days to capture two weekdays and one weekend. Some participants were initially concerned the 3M Tegaderm would cause skin irritation, but we experienced no adverse events. From a research perspective, the devices were easy to set up, extract data, and charge. During the winter session, the wearable sensor was not set up properly for one participant, and so data on posture was not collected.

#### Indoor Positioning System

Six of the 21 participants were willing to use the indoor positioning system. The other 15 participants were not prepared to try the system as they anticipated challenges in the set up, which directly requires connection to their home WiFi router. These participants expressed concerns including not familiar with a home WiFi router, or difficulty accessing the router because it was in a hard-to-reach area. We also learned that four of the 15 participants used a cellular network (Long-Term Evolution system), which was not compatible with our version of the hub design. Overall, the six participants found the indoor positioning system easy to use but provided suggestions to improve the functional design. All six participants reported that the watch was “bulky” and “uncomfortable”. The watch required daily charging and so we ask participants to charge the watch overnight in the room where they slept. Most participants reported few challenges with setting up the beacons or the hub but found the black box design could be improved to be a brighter colour to make the devices less intimidating. Participants were unsure if the beacons and hub were working as there was no indicator light. From a research perspective, linking the data was a little challenging as several participants forgot to calibrate the devices; however, we were able to link it to the other measures. In addition, we know of one participant who switched one beacon to another room mid-way through data collection.

#### Diary

Nine participants reported their daily activities using the ACT24 while the other 12 used a hard copy diary. Initially four participants agreed to use ACT24, but due to challenges in using the software, they decided to complete the hard copy instead. Challenges in using ACT24 included it being “difficult” and “complicated” at first because of the “5-min interval reporting”. Some participants found it challenging to navigate because there were so many options for activities; however, after some practice the majority of participants found ACT24 “fairly easy”. Participants who used the hard copy diary found it easy to complete; four participants had a caregiver complete their hard copy diary. From a research perspective, the hard copy diaries were not a feasible method to collect data as they were not completed with enough detail to extract time, purpose, and social context. The advantage of ACT24 is participants cannot submit an incomplete entry, which encouraged participants to provide enough details about their daily activities. Adherence to the diaries was good with all 20 participants completing either the electronic or hard copy diary probably because they received daily email reminders.

### Adverse Events

We experienced three adverse events among two participants, which were not related to the study. One participant fell twice due to improper footwear or stepping out of the shower onto a damp floor. Another participant with type II diabetes skipped breakfast and felt unwell during the study visit; after consuming orange juice, the person returned to baseline.

## Discussion

We conducted a study in older adults who were pre-frail and frail to understand the feasibility to measure the context of sedentary behaviour in the winter and spring. Context was assessed using a wearable sensor (activPAL4^TM^, posture), a McMaster engineering home monitoring system (indoor positioning system, functional location within the home), and a diary (ACT24 or hard copy diary, purpose and social environment). We met our criteria for recruitment and retention but experienced high refusal rates mainly due to lack of interest or medical mistrust. We found that each measure captured context of sedentary behaviour except for the hard copy diary. Since the hard copy diary was not completed with sufficient detail, we found it challenging to link it to the other two measures. We were able to link data from activPAL4^TM^, the indoor positioning system, and ACT24. All participants were willing to complete the hard copy diary while almost all participants were willing to use the wearable sensor. Only some participants opted to use the indoor positioning system and complete ACT24. The use of wearable sensors, indoor positioning systems, and electronic diaries may be a feasible method to assess context of sedentary behaviour, but more research is needed with device-based measures in diverse groups.

It is unclear what are the best measures to assess context of sedentary behaviour, especially in older adults. Our study is the first study to use a combination of objective and self-report measures to assess context. A recent meta-analysis by Prince et al found that previous studies have used self-report measures (e.g., diaries, questionnaires, surveys) or objective devices (e.g., accelerometers, wearable sensors, pedometers, wearable cameras), but not a combination of both, to assess total sedentary time (12). The most common methods were self-report physical activity and sedentary behaviour questionnaires (single or multi-item questionnaires) or devices including ActiGraph GT3X and activPAL (12). Prince et al found that self-report measures underestimate total sedentary time when compared to device measures by 1.74 hours/day (95% CI -2.11 to -1.38 hours/day) and that single question measures such as the International Physical Activity Questionnaire significantly under-reported sedentary time (12). There is some evidence that multi-item surveys and diaries work reasonably well to assess total sedentary time, particularly when compared to wearable sensors such as activPAL (12,13); however, the meta-analysis included studies mainly conducted in adults between the ages of 18 to 50 years (12). In our study, we found older adults experienced challenges with the hard copy diaries as the diaries were not completed with enough detail. On the other hand, we experienced almost no challenges with activPAL. A 2017 narrative review reported that seven of the nine studies used either ActiGraph or Actiheart accelerometers to assess sedentary behaviour (37); however, accelerometers cannot provide information about posture (e.g., sitting, standing), which is important when defining the context of sedentary behaviour (38). Moreover, wrist-based accelerometers such as ActiGraph are poor estimators of assessing sedentary behaviours and demonstrate the greatest risk for misclassification error (38). For this reason, our study used an wearable sensor (activPAL4^TM^) which can measure time spent sitting, lying, and standing. The advantage of using a thigh-based wearable sensor is it is more accurate at identifying standing postures and movement and can detect transitions from sitting/lying to standing compared to wrist worn devices (38). Although comparisons made with accelerometers show lower accuracy compared to inclinometers, both devices have a great degree of variability within and between studies (up to 6 hours/day of variability with accelerometers and up to 4.6 hours/day with inclinometers) (30). But sedentary behaviour may differ by the season, which was not accounted for in the meta-analysis by Prince and colleagues and may explain the variability between different types of device-based measures. Although our study found that sitting and walking were not different between winter and spring, we had a small sample size and larger studies are needed to determine if there is a difference between seasons. It is probable that one measure alone cannot provide enough information about the context of sedentary behaviour. Future studies need to determine the best combination of devices and self-report measures to assess context of sedentary behaviour specific to older adults, especially those who are frail.

In addition to the wearable sensor and diary, our study utilized a smart home monitoring system with an indoor positioning system to assess functional location of sedentary behaviour within the home. Understanding where older adults spend time sitting during awake hours may be an important factor to consider when developing lifestyle interventions that are unique to that setting or location. Although wearables do not pose any threat to privacy, prolonged use throughout the day may not be practical (39). A 2021 systematic review found barriers to wearables include a relatively short battery life, require maintenance, and cause discomfort over long usage if required to wear consistently throughout the day (39). Participants in our study experienced similar barriers including the smart watch being bulky and uncomfortable to use for long periods and the need to charge the watch daily. Several participants were not willing to configure the smart home system because they were intimidated with the system and set-up process; however, through co-designing processes with end-users, such challenges can easily be overcome. Smart home monitoring systems have several advantages that can be used to monitor health, assess activities of daily living, analyse gait, etc. In addition, smart home systems could be used as a solution to understand context of sedentary behaviour. For example, artificial intelligence, machine learning, and fuzzy logic can be automatically rendered within smart home monitoring systems and be used to identify activities that older adults engage in (e.g., watching TV in the living room). One study developed a robot-integrated smart home (RiSH) for older adults, which used a sensor network to monitor body activities. The RiSH was able to recognize 37 distinct individual activities through sound actions with 88% accuracy and identify falling sounds with 80% accuracy (40). Moreover, smart home systems could also be used to target and decrease certain sedentary behaviours. For example, Rudzicz and colleagues developed a mobile robot to assist older adults with Alzheimer’s disease with their activities of daily living (41); such systems could be used to promote safe mobility among this group. There is a clear advantage to using smart home monitoring systems that utilize artificial intelligence, machine learning, and fuzzy logic that should be piloted with older adults to understand context of sedentary behaviour. However, introducing such technologies also requires educating certain groups that may be mistrustful of the devices. Educational outreach programs and involving diverse groups as patient partners during the co-design process should be conducted in parallel with pilot studies of smart home monitoring systems.

### Strengths and Limitations

Our study had several strengths. We recruited a diverse group of older adults who were mainly frail and had cognitive impairments with diverse demographic characteristics including individuals who only completed grade school or high school. We also used a unique combination of objective and subjective measures to assess context of sedentary behaviour. While our study conformed to the highest standards, our study is not without its limitations. The disadvantage of using only one wearable sensor can result in device failure or corrupt data; we experienced one instance where the data was not captured during the winter period. Although we attempted to recruit diverse individuals (e.g., ethnic minorities, individuals of different genders), we experienced barriers including medical mistrust. Thus, the generalizability of the results may not be feasible in other groups. As this was a feasibility study, we only collected data over three days (two weekdays and one weekend) in the winter and spring, which may not be representative of the season or other time periods (i.e., summer and fall).

## Conclusion

We met our criteria for recruitment and retention but experienced high refusal rates. We recruited 21 older adults who were pre-frail or frail within two months and experienced two dropouts due to medical mistrust or worsening health. We experienced high refusal rates as several participants who initially agreed to participate decided not to enroll. The wearable sensor, indoor positioning system, and ACT24 accurately captured one domain of context but participants experienced challenges completing the hard copy diary. The hard copy was not completed with enough details making it difficult to link it to the other devices. We also found some participants were not willing to utilize the wearable sensor, indoor positioning system, and electronic diary. However, we were able to triage the measures of participants who utilized the wearable sensor, indoor positioning system, and ACT24. Nevertheless, there is some merit to using such devices to capture the context of sedentary behaviour. Future studies will need to determine the most feasible and valid methods to assess the context of sedentary behaviour, especially in diverse older adults.

## Data Availability

Data is publicly available upon request for research purposes only.

## Acknowledgements and funding

The authors would like to thank the following funding agents for their support with the project including McMaster Institute for Research on Aging (MIRA), AGE-WELL, the Hamilton Health Sciences New Investigator Fund, and the Canadian Institutes of Health Research. The corresponding author, who is also the principal investigator, is funded by the 2022 MIRA-AGE-WELL Award and the 2022 CIHR Fellowship Award. We would also like to thank our patient partners, Margaret Denton and Anne Pizzacalla from the Hamilton Council on Aging, and Priscilla Ching from Osteoporosis Canada for their input during the study.

## Author Contributions

IBR conceptualized and designed the study. IBR, RS, ST, AK, JA, and AR assisted with recruitment and IBR, RS, ST, and AK with data collection. QF developed and provided the smart home monitoring system and analyzed and interpreted the data from the system. IBR analyzed, interpreted, and wrote the first draft. All co-authors reviewed the final analysis and revised the original manuscript.

## Other information

This review was registered on clinicaltrials.gov (NCT05661058).

The authors have no competing interests.

Data is publicly available upon request for research purposes only.

## Supporting information

**S1 Table. 2007 Strobe Checklist for Cohort Studies.**

**S1 File. Human Participants Research Checklist.**

**S2 File. Ethics Approval Document.**

**S3 File. TREND Checklist.**

**S4 File. Protocol MAPS-B.**

## Notes

### Competing Interest Statement

The authors have declared no competing interest.

### Funding Statement

IBR received the New Investigator Fund from the Hamilton Health Science (NIF-22541) to support the MAPS-B project. IBR also received funding from the 2022 MIRA-AGE-WELL Award and the 2022 CIHR Fellowship Award. The funders did not play any role in the study design, data collection or analysis, decision to publish, or preparation of the manuscript.

### Author Declarations

Ethics approval was obtained from the Hamilton Integrated Research Ethics Board.

## References

1. Petrusevski C, Choo S, Wilson M, MacDermid J, Richardson J. Interventions to address sedentary behaviour for older adults: a scoping review. Vol. 43, Disability and Rehabilitation. Taylor and Francis Ltd.; 2021. p. 3090–101.

2. Chastin S, Gardiner PA, Harvey JA, Leask CF, Jerez-Roig J, Rosenberg D, et al. Interventions for reducing sedentary behaviour in community-dwelling older adults. Cochrane Database of Systematic Reviews. 2021 Jun 25;2021(6).

3. Walston J, Hadley E, Ferrucci L, Guralnik J, Newman A, Studenski S, et al. Research agenda for frailty in older adults: toward a better understanding of physiology and etiology: summary from the American Geriatrics Society/National Institute on Aging Research Conference on Frailty in Older Adults. J Am Geriatr Soc. 2006;54(6):991–1001.

4. Abellan van Kan G, Rolland YM, Morley JE, Vellas B. Frailty: toward a clinical definition. Vol. 9, Journal of the American Medical Directors Association. United States; 2008. p. 71–2.

5. Owen N, Healy GN, Matthews CE, Dunstan DW. Too much sitting: The population health science of sedentary behavior. Exerc Sport Sci Rev. 2010 Jul;38(3):105–13.

6. Blodgett J, Theou O, Kirkland S, Andreou P, Rockwood K. The association between sedentary behaviour, moderate-vigorousphysical activity and frailty in NHANES cohorts. Maturitas. 2015;80(2):187–91.

7. Katzmarzyk PT, Church TS, Craig CL, Bouchard C. Sitting time and mortality from all causes, cardiovascular disease, and cancer. Med Sci Sports Exerc. 2009 May;41(5):998– 1005.

8. Dogra S, Ashe MC, Biddle SJH, Brown WJ, Buman MP, Chastin S, et al. Sedentary time in older men and women: An international consensus statement and research priorities. Br J Sports Med. 2017 Nov 1;51(21):1526–32.

9. Rawlings GH, Williams RK, Clarke DJ, English C, Fitzsimons C, Holloway I, et al. Exploring adults’ experiences of sedentary behaviour and participation in non-workplace interventions designed to reduce sedentary behaviour: A thematic synthesis of qualitative studies. BMC Public Health. 2019 Aug 13;19(1).

10. Gardiner PA, Clark BK, Healy GN, Eakin EG, Winkler EAH, Owen N. Measuring older adults’ sedentary time: Reliability, validity, and responsiveness. Med Sci Sports Exerc. 2011 Nov;43(11):2127–33.

11. Matthews CE, Chen KY, Freedson PS, Buchowski MS, Beech BM, Pate RR, et al. Amount of time spent in sedentary behaviors in the United States, 2003-2004. Am J Epidemiol. 2008 Apr;167(7):875–81.

12. Prince SA, Cardilli L, Reed JL, Saunders TJ, Kite C, Douillette K, et al. A comparison of self-reported and device measured sedentary behaviour in adults: A systematic review and meta-analysis. Vol. 17, International Journal of Behavioral Nutrition and Physical Activity. BioMed Central Ltd.; 2020.

13. Prince SA, LeBlanc AG, Colley RC, Saunders TJ. Measurement of sedentary behaviour in population health surveys: A review and recommendations. PeerJ. 2017;2017(12).

14. Prince S, LeBlanc A, Colley R, Saunders T. Measurement of sedentary behaviour in population health surveys: a review and recommendations. PeerJ. 2017;5(e4130):5.

15. Chastin S, Schwarz U, Skelton D. Development of a Consensus Taxonomy of Sedentary Behaviors (SIT): Report of Delphi Round 1. PLoS One. 2013;8(12):e82313.

16. Vandenbroucke JP, von Elm E, Altman DG, Gotzsche PC, Mulrow CD, Pocock SJ, et al. Strengthening the Reporting of Observational Studies in Epidemiology (STROBE): Explanation and Elaboration. PLOS Med. 2017;16(4).

17. Tariq S, Kouroukis A, Swance R, Adachi J, Leckie C, Papaioannou A, et al. Recruitment and retention strategies of older adults who are frail for a mobility study. Journal of Clinical Epidemiology.

18. Morley JE, Malmstrom TK, Miller DK. A simple frailty questionnaire (FRAIL) predicts outcomes in middle aged African Americans. J Nutr Health Aging. 2012 Jul;16(7):601–8.

19. Chan CS, Slaughter SE, Allyson Jones C, Ickert C, Wagg AS. Measuring activity performance of older adults using the activpal: A rapid review. Vol. 5, Healthcare (Switzerland). MDPI; 2017.

20. Ianovski A, Fang Q. A Smart Home Platform and Hybrid Indoor Positioning Systems for Enabling Aging in Place [Master’s thesis]. [Hamilton]: McMaster; 2018.

21. Matthews CE, Keadle SK, Moore SC, Schoeller DS, Carroll RJ, Troiano RP, et al. Measurement of active and sedentary behavior in context of large epidemiologic studies. Med Sci Sports Exerc. 2018;50(2):266–76.

22. Matthews CE, Keadle SK, Saint-Maurice PF, Moore SC, Willis EA, Sampson JN, et al. Use of time and energy on exercise, prolonged TV viewing, and work days. Am J Prev Med. 2018;55(3):61–9.

23. Rodrigues IB, Wang E, Keller H, Thabane L, Ashe MC, Brien S, et al. The MoveStrong program for promoting balance and functional strength training and adequate protein intake in pre-frail older adults: A pilot randomized controlled trial. PLoS One. 2021 Sep 1;16(9 September).

24. Nasreddine ZS, Phillips NA, Bédirian V, Charbonneau S, Whitehead V, Collin I, et al. The Montreal Cognitive Assessment, MoCA: A Brief Screening Tool For Mild Cognitive Impairment [Internet]. 2005. Available from: www.mocatest.

25. Kennedy C, Opammodos G, Rockwood K, Thabane L, Adachi J, Kirkland S, et al. A Frailty Index predicts 10-year fracture risk in adults age 25 years and older: results from the Canadian Multicentre Osteoporosis Study (CaMos). Osteoporos Int. 2014;25(12):2825–32.

26. Coventry PA, McMillan D, Clegg A, Brown L, Van Der Feltz-Cornelis C, Gilbody S, et al. Frailty and depression predict instrumental activities of daily living in older adults: A population-based longitudinal study using the CARE75+ cohort. PLoS One. 2020 Dec 1;15(12 December).

27. Xie F, Pullenayegum E, Gaebel K, Bansback N, Bryan S, Ohinmaa A, et al. A Time Trade-off-derived Value Set of the EQ-5D-5L for Canada [Internet]. 2015. Available from: www.lww-medicalcare.com

28. Wancata J, Alexandrowicz R, Marquart B, Weiss M, Friedrich F. The criterion validity of the geriatric depression scale: A systematic review. Vol. 114, Acta Psychiatrica Scandinavica. 2006. p. 398–410.

29. Carlucci L, Balestrieri M, Maso E, Marini A, Conte N, Balsamo M. Psychometric properties and diagnostic accuracy of the short form of the geriatric anxiety scale (GAS-10). BMC Geriatr. 2021 Dec 1;21(1).

30. O’Neill J, Tabish H, Welch V, Petticrew M, Pottie K, Clarke M, et al. Applying an equity lens to interventions: using PROGRESS ensures consideration of socially stratifying factors to illuminate inequities in health. J Clin Epidemiol. 2014;67(1):56–64.

31. Sim J, Lewis M. The size of a pilot study for a clinical trial should be calculated in relation to considerations of precision and efficiency. J Clin Epidemiol [Internet]. 2012;65(3):301–8. Available from: http://dx.doi.org/10.1016/j.jclinepi.2011.07.011

32. Julious SA. Sample size of 12 per group rule of thumb for a pilot study. Pharm Stat. 2005;4:287–91.

33. Eldridge SM, Lancaster GA, Campbell MJ, Thabane L, Hopewell S, Coleman CL, et al. Defining feasibility and pilot studies in preparation for randomised controlled trials: development of a conceptual framework. PLoS One. 2016;11(3).

34. Clegg A, Barber S, Young J, Iliffe S, Forster A. The Home-based Older People’s Exercise (HOPE) trial: A pilot randomised controlled trial of a home-based exercise intervention for older people with frailty. Age Ageing. 2014;43(5):687–95.

35. Provencher V, Mortenson W Ben, Tanguay-Garneau L, Bélanger K, Dagenais M. Challenges and strategies pertaining to recruitment and retention of frail elderly in research studies: A systematic review. Vol. 59, Archives of Gerontology and Geriatrics. Elsevier Ireland Ltd; 2014. p. 18–24.

36. White MD, Marsh EE, White MD, Marsh EE. Content Analysis: A Flexible Methodology Content Analysis: A Flexible Methodology. 2017;55(1):22–45.

37. Copeland JL, Ashe MC, Biddle SJ, Brown WJ, Buman MP, Chastin S, et al. Sedentary time in older adults: A critical review of measurement, associations with health, and interventions. Vol. 51, British Journal of Sports Medicine. BMJ Publishing Group; 2017.

38. Fortune E, Lugade VA, Kaufman KR. Posture and Movement Classification: The Comparison of Tri-Axial Accelerometer Numbers and Anatomical Placement. J Biomech Eng. 2014 May;136(5).

39. Gochoo M, Alnajjar F, Tan TH, Khalid S. Towards privacy-preserved aging in place: A systematic review. Vol. 21, Sensors. MDPI AG; 2021.

40. Do HM, Pham M, Sheng W, Yang D, Liu M. RiSH: A Robot-integrated Smart Home for Elderly Care [Internet]. 2017. Available from: https://www.sciencedirect.com/science/article/pii/S0921889017300477

41. Rudzicz F, Wang R, Begum M, Mihailidis A. Speech interaction with personal assistive robots supporting aging at home for individuals with Alzheimer’s disease. ACM Trans Access Comput. 2015;7(2):1–22.

